# Sleep problems effect on developmental trajectories in children with autism

**DOI:** 10.1101/2022.03.30.22273178

**Authors:** Jonah Levin, Edward Khokhlovich, Andrey Vyshedskiy

**Author notes:** Corresponding author: Andrey Vyshedskiy, Ph.D., Boston University, Boston, USA. Funding: This research received no external funding. Author contributions: AV and EK designed the study. AV and JL analyzed the data. JL and AV wrote the paper. Competing Interests: Authors declare no competing interests.

## Abstract

The effect of sleep problems in 2-to 5-year-old children with ASD was investigated in the largest and the longest observational study to-date. Parents assessed the development of 7069 children quarterly for three years on five orthogonal subscales: receptive language, expressive language, sociability, sensory awareness, and health. Moderate and severe sleep problems were reported in 13% of children. Children with no sleep problems developed faster compared to matched children with sleep problems in all subscales. The greatest difference in trajectories was detected in the health subscale. When controlling for the health score (in addition to each subscale score at baseline as well as gender and severity), the effect of sleep problems decreased in all subscales except the combinatorial receptive language subscale (where the effect of sleep problems was increased), suggesting that sleep problems affect combinatorial language acquisition irrespective of the overall health. This study confirms a high prevalence of sleep problems in ASD children and points to the need for more systematic research as an initial step in developing treatment strategies.

## Introduction

Sleep is well-documented to be crucial for social and behavioral development as well as for mental and physical health. For example, Williams et al. found that sleep problems in 5 to 12 year-old children were associated with emotional and self-regulation problems, hyperactivity, and poorer prosocial skills (Gail Williams et al., 2004). Shimizu et. al. showed that sleep problems in 9 to 11 year-old children predicted greater occurrence of externalizing behaviors, depressive symptoms, and anxiety at the age of 18 years, even when controlling for childhood mental illness (Shimizu et al., 2021). In a review of research on sleep problems in ASD individuals, Cohen et al. found that prevalence of sleep disorders was significantly higher than that in neurotypical children: 40-80% of children with ASD were reported by caregivers to have problems with sleep, as compared to 25-40% in neurotypical children (Cohen et al., 2014). As in neurotypical children, sleep problems in ASD children were reported to be associated with increased severity of behavioral problems and with decrease in prosocial behaviors (Cohen et al., 2014; Hirata et al., 2016; Krakowiak et al., 2008; MacDuffie et al., 2020; Sannar et al., 2018; Schreck et al., 2004), but, in contrast to neurotypical individuals, sleep problems had a tendency to worsen with age in ASD individuals (Deliens & Peigneux, 2019; McLay et al., 2021; Verhoeff et al., 2018).

Sleep problems have also been reported to affect children’s cognitive development and language acquisition. Multiple studies have shown that improved retention of vocabulary, phonological learning, and language comprehension skills are associated with greater amounts of sleep both in neurotypical children (Axelsson et al., 2016; Henderson et al., 2012) and in children with various developmental disorders (Axelsson et al., 2013; Edgin et al., 2015; Fletcher et al., 2020; Greiner de Magalhães et al., 2020; Knowland et al., 2019). Conversely, it has been shown that children who sleep less have more trouble with consolidation of different elements of memory associated with speech, such as phoneme recognition, vocabulary, and semantic knowledge (Axelsson et al., 2016; Bonuck et al., 2021; Henderson et al., 2012). Most studies of cognitive development and language acquisition, however, suffer from low numbers of participants and lack of longitudinal data. In 2015 we published a language training app for children (Dunn, Elgart, Lokshina, Faisman, Khokhlovich, et al., 2017b, 2017a; Dunn, Elgart, Lokshina, Faisman, Waslick, et al., 2017; Vyshedskiy et al., 2020; Vyshedskiy & Dunn, 2015), which invites parents to complete their child’s evaluation and diagnosis every three months. As a result, we accumulated over a hundred thousand longitudinal evaluations. This gives us an opportunity to study children’s developmental trajectories in greater detail. In this report, we investigate the effect of sleep problems reported by parents. To assess the effect of sleep problems, we compared participants with no sleep problems to participants with moderate and serious sleep problems.

## Methods

### Participants

Participants were users of a language therapy app that was made available gratis at all major app stores in September 2015. Once the app was downloaded, caregivers were asked to register and to provide demographic details, including the child’s diagnosis and age. Caregivers consented to anonymized data analysis and completed the Autism Treatment Evaluation Checklist (ATEC) (Rimland & Edelson, 1999), an evaluation of the receptive language using the Mental Synthesis Evaluation Checklist (MSEC) (Braverman et al., 2018), as well as the Screen Time assessment and the Diet and Supplements assessment. The first evaluation was administered approximately one month after the download. The subsequent evaluations were administered at approximately three-month intervals. To enforce regular evaluations, the app became unusable at the end of each three-month interval and parents were required to complete an evaluation to regain its functionality.

#### Inclusion criteria

Inclusion criteria were identical to our previous studies of this population (Fridberg et al., 2021; Mahapatra, Khokhlovich, et al., 2018; Vyshedskiy et al., 2020). Specifically, we selected participants based on the following criteria:

1. Consistency: Participants must have filled out at least three ATEC evaluations and the interval between the first and the last evaluation was six months or longer.
2. Diagnosis: ASD. Children without ASD diagnosis were excluded from the study. Other diagnostic options included: Mild Language Delay, Pervasive Developmental Disorder, Attention Deficit Disorder, Social Communication Disorder, Specific Language Impairment, Apraxia, Sensory Processing Disorder, Down Syndrome, Lost Diagnosis of Autism or PDD, Other Genetic Disorder, normally developed child. The parent-reported ASD diagnosis was supported by ATEC scores. Average initial ATEC total score was 68.5 ± 25.1, which corresponds to moderate-to-severe ASD as delineated earlier (Mahapatra, Vyshedsky, et al., 2018) and Table 1.

**Table 1.**
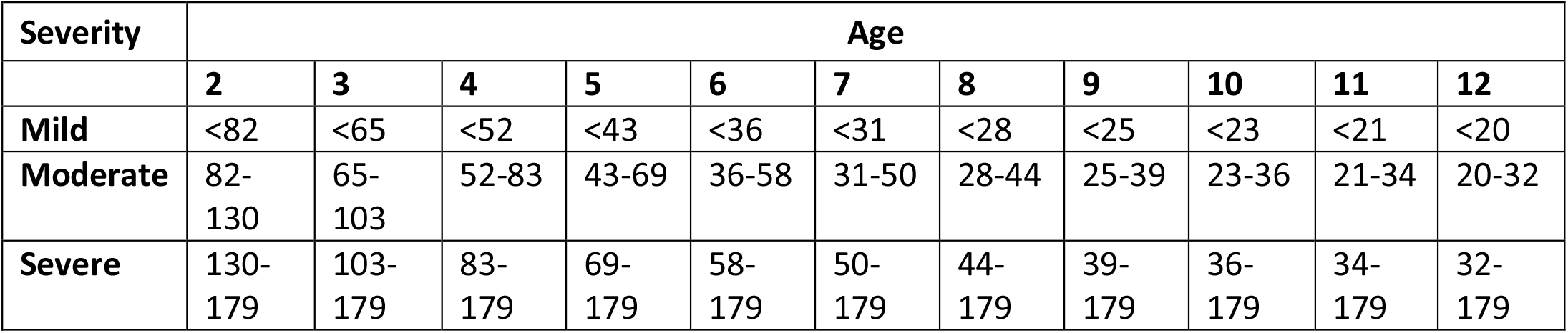
Approximate relationship between ATEC total score, age, and ASD severity as described in Mahapatra *et al*. (Mahapatra, Khokhlovich, et al., 2018). At any age, a greater ATEC score indicates greater ASD severity.

| Severity | Age |  |  |  |  |  |  |  |  |  |  |
| --- | --- | --- | --- | --- | --- | --- | --- | --- | --- | --- | --- |
|  | 2 | 3 | 4 | 5 | 6 | 7 | 8 | 9 | 10 | 11 | 12 |
| **Mild** | <82 | <65 | <52 | <43 | <36 | <31 | <28 | <25 | <23 | <21 | <20 |
| **Moderate** | 82-130 | 65-103 | 52-83 | 43-69 | 36-58 | 31-50 | 28-44 | 25-39 | 23-36 | 21-34 | 20-32 |
| **Severe** | 130-179 | 103-179 | 83-179 | 69-179 | 58-179 | 50-179 | 44-179 | 39-179 | 36-179 | 34-179 | 32-179 |

#### Exclusion criteria

1. Maximum age: Participants older than five years of age at the time of their first evaluation were excluded from this study.
2. Minimum age: Participants who completed their first evaluation before the age of two years were excluded from this study.

After excluding participants that did not meet these criteria, there were 7,069 total participants.

#### Sleep problems assessment

Participants were required to assess sleep problems while responding to the enquiry. The possible answers were: “not a problem,” “minor problem,” “moderate problem,” “serious problem.” To assess the effect of sleep problems, we compared participants with no reported sleep problems (N=4183; the no sleep problems group, abbreviated noSP) to participants with reported moderate and serious sleep problems (N=651; the sleep problems group, abbreviated SP). Participants with minor sleep problems were excluded from analysis. Additionally, participants who reported highly variable sleep problems (standard deviation greater or equal 0.75) were also excluded from the analysis.

### Evaluations

A caregiver-completed Autism Treatment Evaluation Checklist (ATEC) (Rimland & Edelson, 1999) and Mental Synthesis Evaluation Checklist (MSEC) (Braverman et al., 2018) were used to track child development. The ATEC questionnaire is comprised of four subscales: 1) Speech/Language/Communication, 2) Sociability, 3) Sensory/Sensory awareness, and 4) Physical/Health/Behavior. The first subscale, Speech/Language/Communication, contains 14 items and its score ranges from 0 to 28 points. The Sociability subscale contains 20 items within a score range of 0 to 40 points. The third subscale, referred here as the Sensory awareness subscale, has 18 items and score range from 0 to 36 points. The fourth subscale, referred here as the Health subscale, contains 25 items and score range from 0 to 75 points. The scores from each subscale are combined in order to calculate a Total Score, which ranges from 0 to 179 points. A lower score indicates lower severity of ASD symptoms and a higher score indicates more severe symptoms of ASD. ATEC is not a diagnostic checklist and it was designed to evaluate the effectiveness of treatment (Rimland & Edelson, 1999). Therefore, ASD severity can only have an approximate relationship with the total ATEC score and age. Table 1 lists approximate ATEC total score as related to ASD severity and age as described in Mahapatra *et al*. (Mahapatra, Khokhlovich, et al., 2018).

ATEC was selected as a tool since it is one of the few measures validated to evaluate treatment effectiveness. In contrast, another popular ASD assessment tool, Autism Diagnostic Observation Schedule or ADOS, (Lord et al., 2000) has only been validated as a diagnostic tool. Various studies confirmed the validity and reliability of ATEC (Geier et al., 2013; Jarusiewicz, 2002) and several trials confirmed ATEC’s ability to longitudinally measure changes in participant performance (Charman et al., 2004; Klaveness et al., 2013; Magiati et al., 2011; Mahapatra, Khokhlovich, et al., 2018). Moreover, ATEC has been used as a primary outcome measure for a randomized controlled trial of iPad-based intervention for ASD, named “Therapy Outcomes By You” or TOBY, and it was noted that ATEC possesses an “internal consistency and adequate predictive validity” (Whitehouse et al., 2017). These studies support the effectiveness of ATEC as a tool for longitudinal tracking of symptoms and assessing changes in ASD severity.

### Expressive language assessment

The ATEC Speech/Language/Communication subscale includes the following questions: 1) Knows own name, 2) Responds to ‘No’ or ‘Stop’, 3) Can follow some commands, 4) Can use one word at a time (No!, Eat, Water, etc.), 5) Can use 2 words at a time (Don’t want, Go home), 6) Can use 3 words at a time (Want more milk), 7) Knows 10 or more words, 8) Can use sentences with 4 or more words, 9) Explains what he/she wants, 10) Asks meaningful questions, 11) Speech tends to be meaningful/relevant, 12) Often uses several successive sentences, 13) Carries on fairly good conversation, and 14) Has normal ability to communicate for his/her age. With the exception of the first three items, all items in the ATEC subscale 1 primarily measure expressive language. Accordingly, the ATEC subscale 1 is herein referred to as the Expressive Language subscale to distinguish it from the Receptive Language subscale tested by the MSEC evaluation.

### Receptive language assessment

The MSEC evaluation was designed to be complementary to ATEC in measuring complex receptive language. Out of 20 MSEC items, those that directly assess receptive language are the following: 1) Understands simple stories that are read aloud; 2) Understands elaborate fairy tales that are read aloud (i.e., stories describing FANTASY creatures); 3) Understands some simple modifiers (i.e., green apple vs. red apple or big apple vs. small apple); 4) Understands several modifiers in a sentence (i.e., small green apple); 5) Understands size (can select the largest/smallest object out of a collection of objects); 6) Understands possessive pronouns (i.e. your apple vs. her apple); 7) Understands spatial prepositions (i.e., put the apple ON TOP of the box vs. INSIDE the box vs. BEHIND the box); 8) Understands verb tenses (i.e., I will eat an apple vs. I ate an apple); 9) Understands the change in meaning when the order of words is changed (i.e., understands the difference between ‘a cat ate a mouse’ vs. ‘a mouse ate a cat’); 10) Understands explanations about people, objects or situations beyond the immediate surroundings (e.g., “Mom is walking the dog,” “The snow has turned to water”); MSEC consists of 20 questions within a score range of 0 to 40 points; similarly to ATEC, a lower MSEC score indicates a better developed receptive language.

The psychometric quality of MSEC was tested with 3,715 parents of ASD children (Braverman et al., 2018). Internal reliability of MSEC was good (Cronbach’s alpha > 0.9). MSEC exhibited adequate test– retest reliability, good construct validity, and good known group validity as reflected by the difference in MSEC scores for children of different ASD severity levels.

To simplify interpretation of figure labels, the subscale 1 of the ATEC evaluation is herein referred to as the Expressive Language subscale and the MSEC scale is referred to as the Receptive Language subscale.

### Statistical analysis

The framework for the evaluation of score changes over time has been earlier explained in minute detail (Mahapatra, Khokhlovich, et al., 2018; Vyshedskiy et al., 2020). In short, the concept of a “Visit” was developed by dividing the three-year-long observation interval into 3-month periods. All evaluations were mapped into 3-month-long bins with the first evaluation placed in the first bin. When more than one evaluation was completed within a bin, their results were averaged to calculate a single number representing this 3-month interval. Thus, we had 12 quarterly evaluations for both noSP and SP groups.

It was then hypothesized that there was a two-way interaction between pretend-play-group and Visit. Statistically, this hypothesis was modeled by applying the Linear Mixed Effect Model with Repeated Measures (MMRM), where a two-way interaction term was introduced to test the hypothesis. The model (Endpoint ∼ Baseline + Gender + Severity + Sleep-Problem-Group * Visit) was fit using the R Bioconductor library of statistical packages, specifically the “nlme” package. The subscale score at baseline, as well as gender and severity were used as covariates. Conceptually, the model fits a plane into n-dimensional space. This plane considers a complex variability structure across multiple visits, including baseline differences. Once such a plane is fit, the model calculates Least Squares Means (LS Means) for each subscale and group at each visit. The model also calculates LS Mean differences between the groups at each visit.

To control for the health score, a second Linear Mixed Effect Model was used: (Endpoint ∼ Baseline + Gender + Severity + Health + Sleep-Problem-Group * Visit). These results are reported separately.

In preparation for statistical analysis, participants in the SP group were matched to those in the noSP group using propensity score (Schneider et al., 2007) based on age, gender, expressive language, receptive language, sociability, sensory awareness, and health at the 1^st^ evaluation (baseline). The number of matched participants was 643 in each group, Table 2.

**Table 2.** Demographic data of participant pool.

|  | **No sleep problems** | **Sleep problems** |
| --- | --- | --- |
| **Number of participants** | 4,183 | 651 |
| **Number of matched participants** | 643 | 643 |
| **Age at baseline** | 3.5±0.7 | 3.5±0.8 |
| **Male Gender** | 76% | 73% |

## Results

Moderate and severe sleep problems were reported by 13% of participants. These participants were matched to those with no sleep problems using propensity score (Schneider et al., 2007) based on age, gender, expressive language, receptive language, sociability, sensory awareness, and health at the 1^st^ evaluation (baseline). The number of matched participants was 643 out of 651 in the Sleep Problems (SP) group and 643 out of 4183 in the no Sleep Problems (noSP) group. We then analyzed trajectories of children development on five orthogonal subscales: Receptive Language, Expressive Language, Sociability, Sensory awareness, and Health.

On the Receptive Language subscale, the average improvement in the noSP group over 36 months was 8.31 points (SE=0.8, p<0.0001) compared to 6.23 points (SE=1.02, p<0.0001) in the SP group, Figure 1A, Table 3, Table S1. The difference in the noSP group relative to the SP group at Month 36 was not statistically significant: -1.63 points (SE=1.28, p=0.2022). The negative difference (marked in the Table 3 as “noSP – SP”) indicates that the SP group had greater scores at Month 36 and, therefore, more severe symptoms.

**Figure 1.**
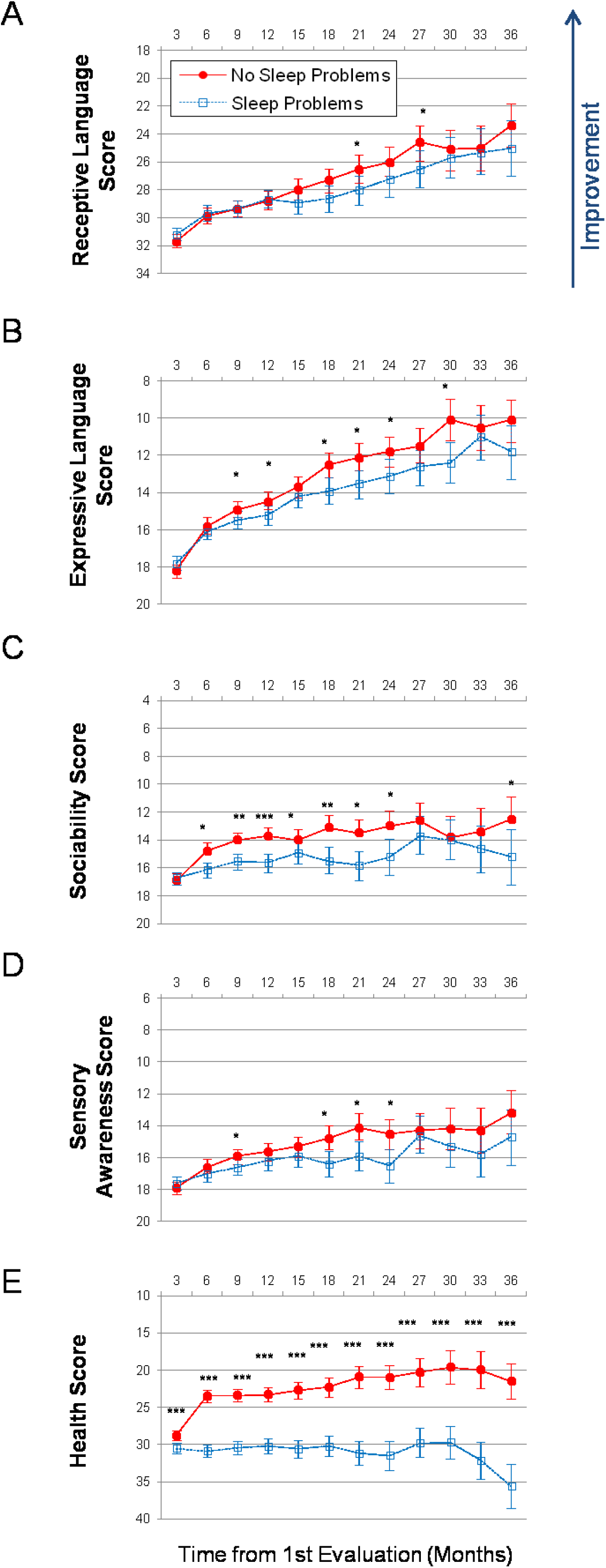
Longitudinal plots of subscale scores LS Means. Horizontal axis shows months from the 1st evaluation (0 to 36 months). Error bars show the 95% confidence interval. To facilitate comparison between subscales, all vertical axes’ ranges have been normalized to show 40% of their corresponding subscale’s maximum available score. A lower score indicates symptom improvement. P-value is marked: ***<0.0001; **<0.001; *<0.05. (A) Receptive Language score. (B) Expressive Language score. (C) Sociability score. (D) Sensory awareness score (E) Health score.

**Table 3.** Characteristics of no Sleep Problems (noSP) and Sleep Problems groups (SP). Data is presented as LS Means (SE; 95% CI). A lower score indicates a lower severity of ASD symptoms. The difference between no Sleep Problems and Sleep Problems (noSP – SP) is presented as: LS Mean (SE; P-value). The negative noSP – SP difference indicates that the SP group had a higher score and therefore more severe symptoms.

| Subscale | Baseline |  |  | Month 36 |  |  | Month 36 – Baseline |  |
| --- | --- | --- | --- | --- | --- | --- | --- | --- |
|  | No Sleep Problems | Sleep Problems | noSP – SP | No Sleep Problems | Sleep Problems | noSP – SP | No Sleep Problems | Sleep Problems |
| **Receptive Language** | 31.7<br>(0.232;<br>31.2 -<br>32.1) | 31.2 (0.245;<br>30.8 - 31.7) | 0.44<br>(0.31;<br>0.1484) | 23.4<br>(0.793;<br>21.8 -<br>24.9) | 25.0<br>(1.015; 23<br>- 27) | -1.63<br>(1.28;<br>0.2022) | -8.31 (0.8;<br><0.0001) | -6.23 (1.02;<br><0.0001) |
| **Expressive Language** | 18.2<br>(0.177;<br>17.89 -<br>18.6) | 17.8 (0.188;<br>17.43 -<br>18.2) | 0.45<br>(0.23;<br>0.0573) | 10.1<br>(0.585; 9 -<br>11.3) | 11.8<br>(0.748;<br>10.3 -<br>13.2) | -1.62<br>(0.94;<br>0.0856) | -8.09<br>(0.59;<br><0.0001) | -6.03 (0.75;<br><0.0001) |
| **Sociability** | 16.9<br>(0.23;<br>16.4 -<br>17.3) | 16.7 (0.242;<br>16.2 - 17.1) | 0.2 (0.31;<br>0.5112) | 10.9<br>(0.541;<br>9.8 - 11.9) | 15.2<br>(1.042;<br>13.2 -<br>17.2) | -2.67<br>(1.31;<br>0.0426) | -4.34<br>(0.82;<br><0.0001) | -1.47 (1.05;<br>0.159) |
| **Sensory awareness** | 17.9<br>(0.197;<br>17.5 -<br>18.3) | 17.6 (0.208;<br>17.2 - 18) | 0.28<br>(0.26;<br>0.2904) | 13.2<br>(0.69;<br>11.8 -<br>14.5) | 14.7<br>(0.883;<br>12.9 -<br>16.4) | -1.51<br>(1.11;<br>0.1751) | -4.73<br>(0.69;<br><0.0001) | -2.95 (0.89;<br>0.0009) |
| **Health** | 28.8<br>(0.342;<br>28.1 -<br>29.4) | 30.5 (0.356;<br>29.8 - 31.2) | -1.75<br>(0.45;<br>0.0001) | 21.5<br>(1.199;<br>19.1 -<br>23.8) | 35.6<br>(1.535;<br>32.6 -<br>38.6) | -14.14<br>(1.93;<br><0.0001) | -7.32<br>(1.21;<br><0.0001) | 5.07 (1.54;<br>0.001) |

On the Expressive Language subscale, the participants in the noSP group improved over the 36-month period by 8.09 points (SE=0.59, p<0.0001) compared to 6.03 points (SE=0.75, p<0.0001) improvement in the SP group, Figure 1B, Table S2. The difference in the noSP group relative to the SP group at Month 36 was: -1.62 points (SE=0.94, p=0.0856).

On the Sociability subscale, the subjects in the noSP group improved over the 36-month period by 4.34 points (SE=0.82, p<0.0001) compared to 1.47 points (SE=1.05, p=0.159) improvement in those in SP group, Figure 1C, Table S3. The difference in the noSP group relative to the SP group at Month 36 was statistically significant: -2.67 points (SE=1.31, p=0.0426).

On the Sensory awareness subscale, the subjects in the noSP group improved over the 36-month period by 4.73 points (SE=0.69, p<0.0001) compared to 2.95 points (SE=0.89, p=0.0009) improvement in the SP group, Figure 1D, Table S4. The difference in the noSP group relative to the SP group at Month 36 was: -1.51 points (SE=1.11, p=0.1751).

On the Health subscale, the subjects in the noSP group improved over the 36-month period by 7.32 points (SE=1.21, p<0.0001) compared to deterioration by 5.07 points (SE=1.54, p=0.001) in the SP group, Figure 1E, Table S5. The difference in the noSP group relative to the SP group at Month 36 was statistically significant: -14.14 points (SE=1.93, p<0.0001).

In order to investigate if the overall health was a driving force behind the group differences, we have re-run the model while controlling for the health score (in addition to each subscale score at baseline as well as gender and severity), Figure 2, Table 4. Controlling for the health score resulted in the increased effect of sleep problems on the combinatorial receptive language subscale and the decreased effect of sleep problems on other subscales. On the combinatorial receptive language subscale the average improvement in the noSP group over 36 months was 8.71 points (SE=0.79, p<0.0001) compared to 5.92 points (SE=0.99, p<0.0001) in the SP group, Table S6. The difference in the noSP group relative to the SP group at Month 36 was statistically significant: -2.54 points (SE=1.26, p<0.0437).

**Figure 2.**
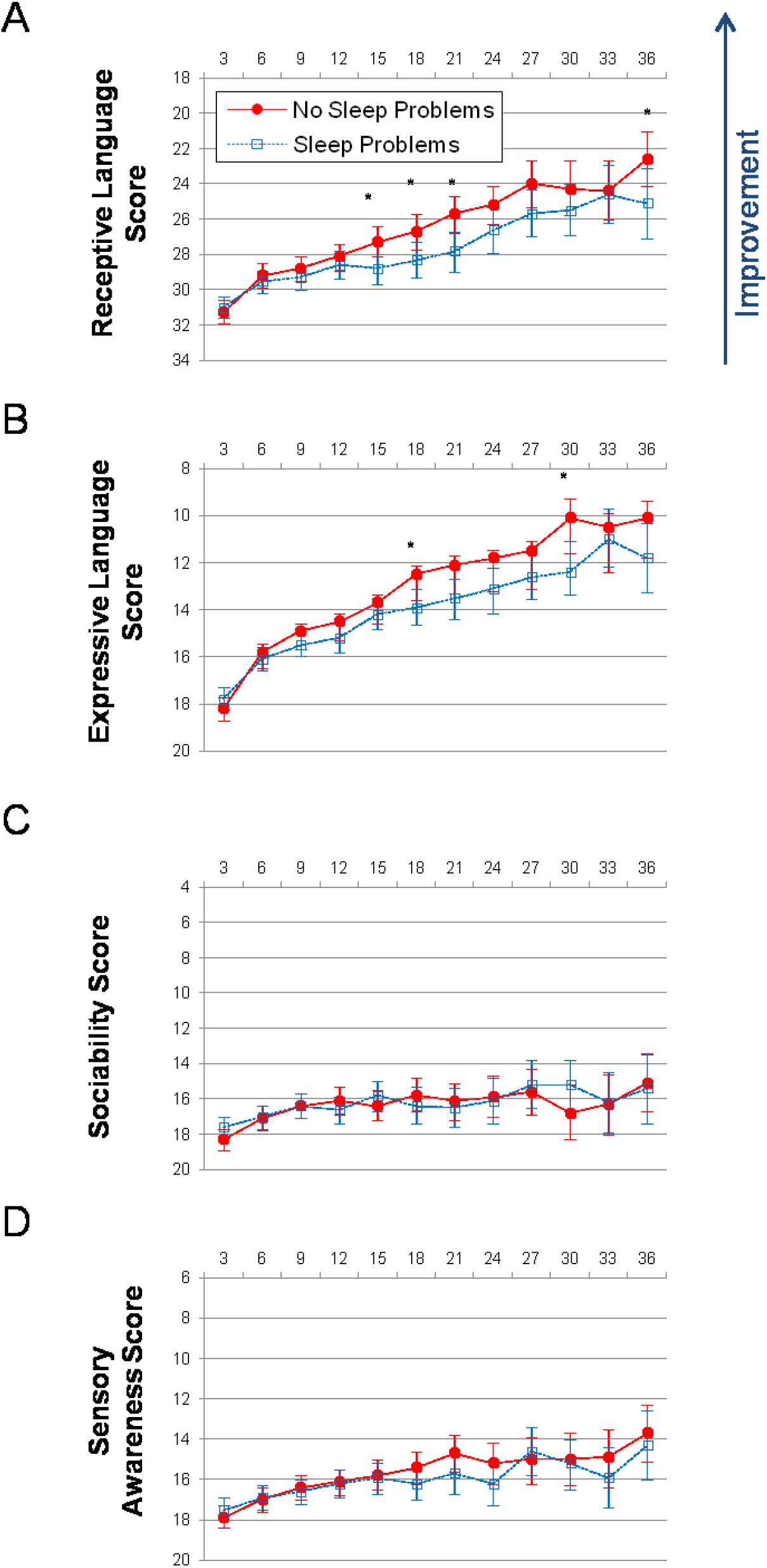
Longitudinal plots of subscale scores LS Means when controlling for the health score. Horizontal axis shows months from the 1st evaluation (0 to 36 months). Error bars show the 95% confidence interval. To facilitate comparison between subscales, all vertical axes ranges have been normalized to show 40% of their corresponding subscale’s maximum available score. A lower score indicates symptom improvement. P-value is marked: ***<0.0001; **<0.001; *<0.05. (A) Receptive Language score. (B) Expressive Language score. (C) Sociability score. (D) Sensory awareness score.

**Table 4.**
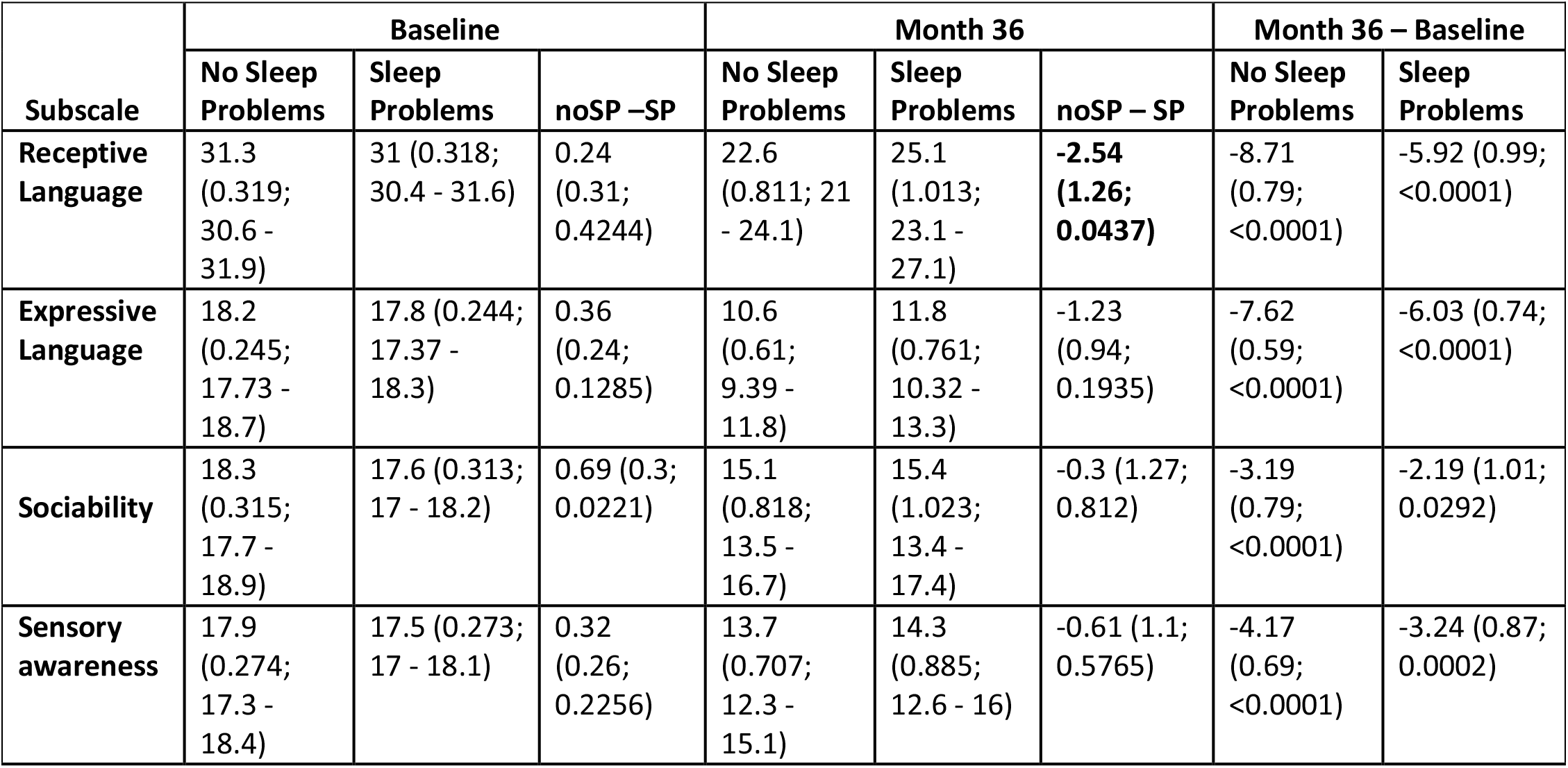
Characteristics of no Sleep Problems (noSP) and Sleep Problems groups (SP) when controlling for the health score. Data is presented as LS Means (SE; 95% CI). A lower score indicates a lower severity of ASD symptoms. The difference between no Sleep Problems and Sleep Problems (noSP – SP) is presented as: LS Mean (SE; P-value). The negative noSP – SP difference indicates that the SP group had a higher score and therefore more severe symptoms.

| Subscale | Baseline |  |  | Month 36 |  |  | Month 36 – Baseline |  |
| --- | --- | --- | --- | --- | --- | --- | --- | --- |
|  | No Sleep Problems | Sleep Problems | noSP – SP | No Sleep Problems | Sleep Problems | noSP – SP | No Sleep Problems | Sleep Problems |
| **Receptive Language** | 31.3 (0.319; 30.6 - 31.9) | 31 (0.318; 30.4 - 31.6) | 0.24 (0.31; 0.4244) | 22.6 (0.811; 21 - 24.1) | 25.1 (1.013; 23.1 - 27.1) | **-2.54 (1.26; 0.0437)** | -8.71 (0.79; <0.0001) | -5.92 (0.99; <0.0001) |
| **Expressive Language** | 18.2 (0.245; 17.73 - 18.7) | 17.8 (0.244; 17.37 - 18.3) | 0.36 (0.24; 0.1285) | 10.6 (0.61; 9.39 - 11.8) | 11.8 (0.761; 10.32 - 13.3) | -1.23 (0.94; 0.1935) | -7.62 (0.59; <0.0001) | -6.03 (0.74; <0.0001) |
| **Sociability** | 18.3 (0.315; 17.7 - 18.9) | 17.6 (0.313; 17 - 18.2) | 0.69 (0.3; 0.0221) | 15.1 (0.818; 13.5 - 16.7) | 15.4 (1.023; 13.4 - 17.4) | -0.3 (1.27; 0.812) | -3.19 (0.79; <0.0001) | -2.19 (1.01; 0.0292) |
| **Sensory awareness** | 17.9 (0.274; 17.3 - 18.4) | 17.5 (0.273; 17 - 18.1) | 0.32 (0.26; 0.2256) | 13.7 (0.707; 12.3 - 15.1) | 14.3 (0.885; 12.6 - 16) | -0.61 (1.1; 0.5765) | -4.17 (0.69; <0.0001) | -3.24 (0.87; 0.0002) |

The effect of sleep problems on expressive language decreased when controlling for the health score (in addition to the expressive language score at baseline as well as gender and severity). The noSP group improved over the 36-month period by 7.62 points (SE=0.59, p<0.0001) compared to 6.03 points (SE=0.74, p<0.0001) improvement in the SP group, Table S7. The difference in the noSP group relative to SP group at Month 36 was: -1.23 points (SE=0.94, p=0.1935).

The effect of sleep problems on sociability decreased when controlling for the health score (in addition to the sociability score at baseline as well as gender and severity). The noSP group improved over the 36-month period by 3.19 points (SE=0.79, p<0.0001) compared to 2.19-point improvement in the SP group (SE=1.01, p=0.0292), Table S8. The difference in the noSP group relative to SP group at Month 36 was: -0.3 points (SE=1.27, p=0.812).

The effect of sleep problems on sensory awareness also decreased when controlling for the health score (in addition to the sensory awareness score at baseline as well as gender and severity). The noSP group improved over the 36-month period by 4.17 points (SE=0.69, p<0.0001) compared to 3.24 point improvement in the SP group (SE=0.87, p=0.0002), Table S9. The difference in the noSP group relative to SP group at Month 36 was: -0.61 points (SE=1.1, p=0.5765).

## Discussion

This is the largest study to date demonstrating a strong association between sleep problems and development in children. Parents assessed the development of 7069 two-to five-year-old children with ASD quarterly for three years. The no Sleep Problems (noSP) group included 4,183 children with no sleep problems and the Sleep Problems (SP) group included 651 children with moderate and severe sleep problems. In order to compare the groups, participants in the SP group were matched to those in the noSP group using propensity score (Schneider et al., 2007) based on age, gender, expressive language, receptive language, sociability, sensory awareness, and health at the 1^st^ evaluation (baseline). The number of matched participants was 643 in each group.

Participants in the no Sleep Problems (noSP) group improved to a greater extent than matched participants in the Sleep Problems (SP) group in all subscales. The difference reached statistical significance in the Combinatorial Receptive Language subscale on Months 21 and 27; in the Expressive Language subscale on Months 9 through 24 and Month 30; in the Sociability subscale on Months 6 through 24 and Month 36; in the Sensory awareness subscale on Months 9, 18, 21, 24; in the Health subscale on all time points. When controlling for the health score (in addition to each subscale score at baseline as well as gender and severity), the effect of sleep problems increased on the combinatorial receptive language subscale and decreased in other subscales, suggesting that sleep problems affect cognitive function irrespective of the overall health. The difference reached statistical significance in the Combinatorial Receptive Language subscale on Months 15, 18, 21 and 36; in the Expressive Language subscale on Months 18 and 30; and never on the Sociability and the Sensory awareness subscales. The results of this study support previous reports of a negative correlation between the presence of sleep problems and every aspect of a child’s development.

These findings add significantly to the results of previous studies due to the longitudinal nature of the collected data. There is almost no longitudinal data looking at sleep problems in children with ASD, with the vast majority of studies being cross-sectional. Looking at the changes in the score over three years provides a growing pool of evidence to what was previously primarily a conjecture. Going forward, increased longitudinal studies like this will continue to add new information about the development of children with ASD who have different types of sleep problems.

This data also adds to the research of how language acquisition is affected by sleep problems in ASD. Few studies focused on this topic and all of them used vocabulary to assess the effect of sleep problems on language (Botting & Baraka, 2018; Fletcher et al., 2020; Knowland et al., 2019). Going forward, one would suggest shifting focus toward receptive and expressive language. This, in turn, could provide further data with respect to the longitudinal development of a wider array of language, cognitive, and communicative skills and their connection to sleep abnormalities in children with ASD.

Interestingly, the proportion of children with ASD suffering from sleep problems in this study (13%) was smaller than those previously reported (40-80%) (Cohen et al., 2014). This, however, is likely due to the fact that children with minor sleep problems were not included in the SP group, whereas previous studies included moderate, severe, and minor sleep problems in their data.

## Data Availability

All data produced in the present study are available upon reasonable request to the authors

## Funding

This research did not receive any specific grant from funding agencies in the public, commercial, or not-for-profit sectors.

## Acknowledgements

We wish to thank Dr. Petr Ilyinskii for his scrupulous editing of this manuscript.

## Author contributions

AV and EK designed the study. AV and JL analyzed the data. AV and JL wrote the paper.

## Competing Interests

Authors declare no competing interests.

## Informed Consent

Caregivers have consented to anonymized data analysis and publication of the results. The study was conducted in compliance with the Declaration of Helsinki (Association, 2013).

## Compliance with Ethical Standards

Using the Department of Health and Human Services regulations found at 45 CFR 46.101(b)(4), the Biomedical Research Alliance of New York LLC Institutional Review Board (IRB) determined that this research project is exempt from IRB oversight.

## Data Availability

De-identified raw data from this manuscript are available from the corresponding author upon reasonable request.

## Code availability statement

Code is available from the corresponding author upon reasonable request.

## Notes

### Competing Interest Statement

The authors have declared no competing interest.

### Funding Statement

This study did not receive any funding

## References

Association, W. M. (2013). World Medical Association Declaration of Helsinki: Ethical principles for medical research involving human subjects. Jama, 310(20), 2191–2194.

Axelsson, E. L., Hill, C. M., Sadeh, A., & Dimitriou, D. (2013). Sleep problems and language development in toddlers with Williams syndrome. Research in Developmental Disabilities, 34(11), 3988–3996.

Axelsson, E. L., Williams, S. E., & Horst, J. S. (2016). The effect of sleep on children’s word retention and generalization. Frontiers in Psychology, 7, 1192.

Bonuck, K., Battino, R., Barresi, I., & McGrath, K. (2021). Sleep problem screening of young children by speech-language pathologists: A mixed-methods feasibility study. Autism & Developmental Language Impairments, 6, 23969415211035064.

Botting, N., & Baraka, N. (2018). Sleep behaviour relates to language skills in children with and without communication disorders. International Journal of Developmental Disabilities, 64(4–5), 238–243.

Braverman, J., Dunn, R., & Vyshedskiy, A. (2018). Development of the Mental Synthesis Evaluation Checklist (MSEC): A Parent-Report Tool for Mental Synthesis Ability Assessment in Children with Language Delay. Children, 5(5), 62. https://doi.org/10.3390/children5050062

Charman, T., Howlin, P., Berry, B., & Prince, E. (2004). Measuring developmental progress of children with autism spectrum disorder on school entry using parent report. Autism, 8(1), 89–100.

Cohen, S., Conduit, R., Lockley, S. W., Rajaratnam, S. M., & Cornish, K. M. (2014). The relationship between sleep and behavior in autism spectrum disorder (ASD): A review. Journal of Neurodevelopmental Disorders, 6(1), 1–10.

Deliens, G., & Peigneux, P. (2019). Sleep–behaviour relationship in children with autism spectrum disorder: Methodological pitfalls and insights from cognition and sensory processing. Developmental Medicine & Child Neurology, 61(12), 1368–1376.

Dunn, R., Elgart, J., Lokshina, L., Faisman, A., Khokhlovich, E., Gankin, Y., & Vyshedskiy, A. (2017a). Children With Autism Appear To Benefit From Parent-Administered Computerized Cognitive And Language Exercises Independent Of the Child’s Age Or Autism Severity. Autism Open Access, 7(217). https://doi.org/10.4172/2165-7890.1000217

Dunn, R., Elgart, J., Lokshina, L., Faisman, A., Khokhlovich, E., Gankin, Y., & Vyshedskiy, A. (2017b). Comparison of performance on verbal and nonverbal multiple-cue responding tasks in children with ASD. Autism Open Access, 7, 218. https://doi.org/10.4172/2165-7890.1000218

Dunn, R., Elgart, J., Lokshina, L., Faisman, A., Waslick, M., Gankin, Y., & Vyshedskiy, A. (2017). Tablet-Based Cognitive Exercises as an Early Parent-Administered Intervention Tool for Toddlers with Autism—Evidence from a Field Study. Clinical Psychiatry, 3(1). https://doi.org/10.21767/2471-9854.100037

Edgin, J. O., Tooley, U., Demara, B., Nyhuis, C., Anand, P., & Spanò, G. (2015). Sleep disturbance and expressive language development in preschool-age children with Down syndrome. Child Development, 86(6), 1984–1998.

Fletcher, F. E., Knowland, V., Walker, S., Gaskell, M. G., Norbury, C., & Henderson, L. M. (2020). Atypicalities in sleep and semantic consolidation in autism. Developmental Science, 23(3), e12906.

Fridberg, E., Khokhlovich, E., & Vyshedskiy, A. (2021). Watching Videos and Television Is Related to a Lower Development of Complex Language Comprehension in Young Children with Autism. Healthcare, 9(4), 423.

Gail Williams, P., Sears, L. L., & Allard, A. (2004). Sleep problems in children with autism. Journal of Sleep Research, 13(3), 265–268.

Geier, D. A., Kern, J. K., & Geier, M. R. (2013). A comparison of the Autism Treatment Evaluation Checklist (ATEC) and the Childhood Autism Rating Scale (CARS) for the quantitative evaluation of autism. Journal of Mental Health Research in Intellectual Disabilities, 6(4), 255–267.

Greiner de Magalhães, C., O’Brien, L. M., & Mervis, C. B. (2020). Sleep characteristics and problems of 2-year-olds with Williams syndrome: Relations with language and behavior. Journal of Neurodevelopmental Disorders, 12(1), 1–16.

Henderson, L. M., Weighall, A. R., Brown, H., & Gareth Gaskell, M. (2012). Consolidation of vocabulary is associated with sleep in children. Developmental Science, 15(5), 674–687.

Hirata, I., Mohri, I., Kato-Nishimura, K., Tachibana, M., Kuwada, A., Kagitani-Shimono, K., Ohno, Y., Ozono, K., & Taniike, M. (2016). Sleep problems are more frequent and associated with problematic behaviors in preschoolers with autism spectrum disorder. Research in Developmental Disabilities, 49, 86–99.

Jarusiewicz, B. (2002). Efficacy of Neurofeedback for Children in the Autistic Spectrum: A Pilot Study. Journal of Neurotherapy, 6(4), 39–49. https://doi.org/10.1300/J184v06n04_05

Klaveness, J., Bigam, J., & Reichelt, K. L. (2013). The varied rate of response to dietary intervention in autistic children. Open Journal of Psychiatry, 3(02), 56.

Knowland, V. C., Fletcher, F., Henderson, L.-M., Walker, S., Norbury, C. F., & Gaskell, M. G. (2019). Sleep promotes phonological learning in children across language and autism spectra. Journal of Speech, Language, and Hearing Research, 62(12), 4235–4255.

Krakowiak, P., Goodlin-Jones, B., Hertz-Picciotto, I., Croen, L. A., & Hansen, R. L. (2008). Sleep problems in children with autism spectrum disorders, developmental delays, and typical development: A population-based study. Journal of Sleep Research, 17(2), 197–206.

Lord, C., Risi, S., Lambrecht, L., Cook, E. H., Leventhal, B. L., DiLavore, P. C., Pickles, A., & Rutter, M. (2000). The Autism Diagnostic Observation Schedule—Generic: A standard measure of social and communication deficits associated with the spectrum of autism. Journal of Autism and Developmental Disorders, 30(3), 205–223.

MacDuffie, K. E., Munson, J., Greenson, J., Ward, T. M., Rogers, S. J., Dawson, G., & Estes, A. (2020). Sleep problems and trajectories of restricted and repetitive behaviors in children with neurodevelopmental disabilities. Journal of Autism and Developmental Disorders, 50(11), 3844–3856.

Magiati, I., Moss, J., Yates, R., Charman, T., & Howlin, P. (2011). Is the Autism Treatment Evaluation Checklist a useful tool for monitoring progress in children with autism spectrum disorders? Journal of Intellectual Disability Research, 55(3), 302–312. https://doi.org/10.1111/j.1365-2788.2010.01359.x

Mahapatra, S., Khokhlovich, E., Martinez, S., Kannel, B., Edelson, S. M., & Vyshedskiy, A. (2018). Longitudinal Epidemiological Study of Autism Subgroups Using Autism Treatment Evaluation Checklist (ATEC) Score. Autism and Developmental Disorders, 1(12). https://doi.org/10.1007/s10803-018-3699-2

Mahapatra, S., Vyshedsky, D., Martinez, S., Kannel, B., Braverman, J., Edelson, S. M., & Vyshedskiy, A. (2018). Autism Treatment Evaluation Checklist (ATEC) norms: A “growth chart” for ATEC score changes as a function of age. Children, 5(2). https://doi.org/10.3390

McLay, L., France, K., Blampied, N., van Deurs, J., Hunter, J., Knight, J., Hastie, B., Carnett, A., Woodford, E., & Gibbs, R. (2021). Function-based behavioral interventions for sleep problems in children and adolescents with autism: Summary of 41 clinical cases. Journal of Autism and Developmental Disorders, 51(2), 418–432.

Rimland, B., & Edelson, S. (1999). Autism Research Institute. Autism Treatment Evaluation Checklist (ATEC).

Sannar, E. M., Palka, T., Beresford, C., Peura, C., Kaplan, D., Verdi, M., Siegel, M., Kaplan, S., & Grados, M. (2018). Sleep problems and their relationship to maladaptive behavior severity in psychiatrically hospitalized children with autism spectrum disorder (ASD). Journal of Autism and Developmental Disorders, 48(11), 3720–3726.

Schneider, B., Carnoy, M., Kilpatrick, J., Schmidt, W. H., & Shavelson, R. J. (2007). Estimating causal effects using experimental and observational design. American Educational & Reseach Association.

Schreck, K. A., Mulick, J. A., & Smith, A. F. (2004). Sleep problems as possible predictors of intensified symptoms of autism. Research in Developmental Disabilities, 25(1), 57–66.

Shimizu, M., Zeringue, M. M., Erath, S. A., Hinnant, J. B., & El-Sheikh, M. (2021). Trajectories of sleep problems in childhood: Associations with mental health in adolescence. Sleep, 44(3), zsaa190.

Verhoeff, M. E., Blanken, L. M., Kocevska, D., Mileva-Seitz, V. R., Jaddoe, V. W., White, T., Verhulst, F., Luijk, M. P., & Tiemeier, H. (2018). The bidirectional association between sleep problems and autism spectrum disorder: A population-based cohort study. Molecular Autism, 9(1), 1–9.

Vyshedskiy, A., & Dunn, R. (2015). Mental Imagery Therapy for Autism (MITA)-An Early Intervention Computerized Brain Training Program for Children with ASD. Autism Open Access, 5(1000153), 2.

Vyshedskiy, A., Khokhlovich, E., Dunn, R., Faisman, A., Elgart, J., Lokshina, L., Gankin, Y., Ostrovsky, S., deTorres, L., & Edelson, S. M. (2020). Novel prefrontal synthesis intervention improves language in children with autism. Healthcare, 8(4), 566. https://doi.org/doi.org/10.3390/healthcare8040566

Whitehouse, A. J., Granich, J., Alvares, G., Busacca, M., Cooper, M. N., Dass, A., Duong, T., Harper, R., Marshall, W., Richdale, A., & others. (2017). A randomised controlled trial of an iPad-based application to complement early behavioural intervention in Autism Spectrum Disorder. Journal of Child Psychology and Psychiatry, 58(9), 1042–1052.

