## Supplementary Material for "Sleep problems effect on developmental trajectories in children with autism"

**Table S1: LS Means (SE; 95% CI) for Receptive Language MSEC subscale score. The difference between no Sleep Problems and Sleep Problems (noSP – SP) is presented as: LS Mean (SE; P-value). The negative noSP – SP difference indicates that the SP group had higher score and therefore more severe symptoms.**

| **Visit Number** | **No Sleep Problems group** | **Sleep Problems group** | **noSP – SP** |
| --- | --- | --- | --- |
| Baseline | 31.7 (0.232; 31.2 - 32.1) | 31.2 (0.245; 30.8 - 31.7) | 0.44 (0.31; 0.1484) |
| Month 6 | 29.9 (0.279; 29.3 - 30.4) | 29.7 (0.292; 29.2 - 30.3) | 0.16 (0.38; 0.6832) |
| Month 9 | 29.4 (0.28; 28.8 - 29.9) | 29.4 (0.299; 28.8 - 30) | -0.05 (0.39; 0.9009) |
| Month 12 | 28.8 (0.323; 28.1 - 29.4) | 28.7 (0.344; 28.1 - 29.4) | 0.02 (0.45; 0.971) |
| Month 15 | 28 (0.382; 27.2 - 28.7) | 28.9 (0.4; 28.1 - 29.7) | -0.97 (0.53; 0.0696) |
| Month 18 | 27.3 (0.445; 26.5 - 28.2) | 28.6 (0.477; 27.6 - 29.5) | -1.22 (0.64; 0.0541) |
| Month 21 | 26.5 (0.491; 25.5 - 27.5) | 28 (0.537; 26.9 - 29) | **-1.47 (0.71; 0.0397)** |
| Month 24 | 26 (0.539; 24.9 - 27) | 27.2 (0.64; 25.9 - 28.5) | -1.21 (0.82; 0.1396) |
| Month 27 | 24.6 (0.64; 23.4 - 25.9) | 26.5 (0.671; 25.2 - 27.8) | **-1.86 (0.92; 0.0425)** |
| Month 30 | 25.1 (0.753; 23.7 - 26.6) | 25.7 (0.738; 24.3 - 27.2) | -0.6 (1.04; 0.5645) |
| Month 33 | 25 (0.826; 23.4 - 26.6) | 25.3 (0.843; 23.7 - 27) | -0.33 (1.17; 0.777) |
| Month 36 | 23.4 (0.793; 21.8 - 24.9) | 25.0 (1.015; 23 - 27) | -1.63 (1.28; 0.2022) |
| Month 36 - Baseline | -8.31 (0.8; <0.0001) | -6.23 (1.02; <0.0001) | na |

**Table S2: LS Means (SE; 95% CI) for Expressive Language measured by the Subscale 1 of ATEC. The difference between no Sleep Problems and Sleep Problems (noSP – SP) is presented as: LS Mean (SE; P-value). The negative noSP – SP difference indicates that the SP group had higher score and therefore more severe symptoms.**

| **Visit Number** | **No Sleep Problems group** | **Sleep Problems group** | **noSP – SP** |
| --- | --- | --- | --- |
| Baseline | 18.2 (0.177; 17.89 - 18.6) | 17.8 (0.188; 17.43 - 18.2) | 0.45 (0.23; 0.0573) |
| Month 6 | 15.8 (0.211; 15.34 - 16.2) | 16.1 (0.221; 15.68 - 16.5) | -0.36 (0.29; 0.2143) |
| Month 9 | 14.9 (0.212; 14.47 - 15.3) | 15.5 (0.226; 15.08 - 16) | **-0.64 (0.29; 0.0283)** |
| Month 12 | 14.5 (0.243; 13.97 - 14.9) | 15.2 (0.259; 14.65 - 15.7) | **-0.71 (0.34; 0.0362)** |
| Month 15 | 13.7 (0.286; 13.15 - 14.3) | 14.2 (0.299; 13.6 - 14.8) | -0.48 (0.4; 0.2325) |
| Month 18 | 12.5 (0.331; 11.85 - 13.2) | 13.9 (0.355; 13.17 - 14.6) | **-1.36 (0.47; 0.0039)** |
| Month 21 | 12.1 (0.364; 11.35 - 12.8) | 13.5 (0.399; 12.67 - 14.2) | **-1.38 (0.53; 0.0087)** |
| Month 24 | 11.8 (0.399; 11.01 - 12.6) | 13.1 (0.473; 12.14 - 14) | **-1.28 (0.61; 0.0349)** |
| Month 27 | 11.5 (0.473; 10.53 - 12.4) | 12.6 (0.496; 11.6 - 13.5) | -1.11 (0.68; 0.1008) |
| Month 30 | 10.1 (0.555; 8.97 - 11.2) | 12.4 (0.546; 11.33 - 13.5) | **-2.34 (0.77; 0.0024)** |
| Month 33 | 10.5 (0.609; 9.31 - 11.7) | 11 (0.623; 9.76 - 12.2) | -0.48 (0.86; 0.5797) |
| Month 36 | 10.1 (0.585; 9 - 11.3) | 11.8 (0.748; 10.3 - 13.2) | -1.62 (0.94; 0.0856) |
| Month 36 - Baseline | -8.09 (0.59; <0.0001) | -6.03 (0.75; <0.0001) | na |

**Table S3: LS Means (SE; 95% CI) for Sociability subscale score measured by the Subscale 2 of ATEC. The difference between no Sleep Problems and Sleep Problems (noSP – SP) is presented as: LS Mean (SE; P-value). The negative noSP – SP difference indicates that the SP group had higher score and therefore more severe symptoms.**

| **Visit Number** | **No Sleep Problems group** | **Sleep Problems group** | **noSP – SP** |
| --- | --- | --- | --- |
| Baseline | 16.9 (0.23; 16.4 - 17.3) | 16.7 (0.242; 16.2 - 17.1) | 0.2 (0.31; 0.5112) |
| Month 6 | 14.8 (0.28; 14.2 - 15.3) | 16.1 (0.291; 15.5 - 16.6) | **-1.26 (0.38; 0.001)** |
| Month 9 | 14 (0.28; 13.5 - 14.6) | 15.5 (0.299; 14.9 - 16) | **-1.45 (0.39; 0.0002)** |
| Month 12 | 13.7 (0.326; 13.1 - 14.3) | 15.6 (0.346; 14.9 - 16.2) | **-1.87 (0.45; <0.0001)** |
| Month 15 | 14 (0.388; 13.2 - 14.7) | 14.9 (0.405; 14.1 - 15.7) | **-0.92 (0.54; 0.0919)** |
| Month 18 | 13.1 (0.452; 12.2 - 14) | 15.5 (0.484; 14.6 - 16.5) | **-2.37 (0.65; 0.0003)** |
| Month 21 | 13.5 (0.5; 12.5 - 14.4) | 15.8 (0.548; 14.7 - 16.8) | **-2.3 (0.73; 0.0016)** |
| Month 24 | 13 (0.551; 11.9 - 14) | 15.2 (0.653; 13.9 - 16.5) | **-2.22 (0.84; 0.0084)** |
| Month 27 | 12.6 (0.656; 11.3 - 13.9) | 13.7 (0.686; 12.4 - 15.1) | -1.11 (0.94; 0.2357) |
| Month 30 | 13.8 (0.772; 12.3 - 15.3) | 14 (0.755; 12.6 - 15.5) | -0.26 (1.07; 0.8097) |
| Month 33 | 13.4 (0.847; 11.7 - 15) | 14.6 (0.864; 12.9 - 16.2) | -1.17 (1.2; 0.3291) |
| Month 36 | 12.5 (0.814; 10.9 - 14.1) | 15.2 (1.042; 13.2 - 17.2) | **-2.67 (1.31; 0.0426)** |
| Month 36 - Baseline | -4.34 (0.82; <0.0001) | -1.47 (1.05; 0.159) | na |

**Table S4: LS Means (SE; 95% CI) for the Sensory/Cognitive Awareness subscale score measured by the Subscale 3 of ATEC. The difference between no Sleep Problems and Sleep Problems (noSP – SP) is presented as: LS Mean (SE; P-value). The negative noSP – SP difference indicates that the SP group had higher score and therefore more severe symptoms.**

| **Visit Number** | **No Sleep Problems group** | **Sleep Problems group** | **noSP – SP** |
| --- | --- | --- | --- |
| Baseline | 17.9 (0.197; 17.5 - 18.3) | 17.6 (0.208; 17.2 - 18) | 0.28 (0.26; 0.2904) |
| Month 6 | 16.6 (0.239; 16.1 - 17) | 17 (0.249; 16.5 - 17.5) | -0.45 (0.32; 0.1687) |
| Month 9 | 15.9 (0.239; 15.5 - 16.4) | 16.6 (0.256; 16.1 - 17.1) | **-0.69 (0.33; 0.0375)** |
| Month 12 | 15.6 (0.278; 15.1 - 16.2) | 16.2 (0.296; 15.6 - 16.8) | -0.61 (0.39; 0.1168) |
| Month 15 | 15.3 (0.329; 14.7 - 16) | 15.9 (0.345; 15.2 - 16.6) | -0.6 (0.46; 0.1926) |
| Month 18 | 14.8 (0.384; 14 - 15.5) | 16.4 (0.412; 15.6 - 17.2) | **-1.61 (0.55; 0.0034)** |
| Month 21 | 14.1 (0.425; 13.2 - 14.9) | 15.9 (0.466; 15 - 16.8) | **-1.86 (0.62; 0.0025)** |
| Month 24 | 14.5 (0.467; 13.6 - 15.5) | 16.5 (0.555; 15.4 - 17.5) | **-1.91 (0.71; 0.0073)** |
| Month 27 | 14.3 (0.556; 13.2 - 15.4) | 14.6 (0.583; 13.5 - 15.8) | -0.38 (0.79; 0.6365) |
| Month 30 | 14.2 (0.654; 12.9 - 15.5) | 15.3 (0.641; 14 - 16.5) | -1.02 (0.91; 0.2586) |
| Month 33 | 14.3 (0.718; 12.9 - 15.7) | 15.8 (0.734; 14.4 - 17.2) | -1.54 (1.02; 0.1315) |
| Month 36 | 13.2 (0.69; 11.8 - 14.5) | 14.7 (0.883; 12.9 - 16.4) | -1.51 (1.11; 0.1751) |
| Month 36 - Baseline | -4.73 (0.69; <0.0001) | -2.95 (0.89; 0.0009) | na |

**Table S5: LS Means (SE; 95% CI) for Health/Physical/Behavior subscale score measured by the Subscale 4 of ATEC. The difference between no Sleep Problems and Sleep Problems (noSP – SP) is presented as: LS Mean (SE; P-value). The negative noSP – SP difference indicates that the SP group had higher score and therefore more severe symptoms.**

| **Visit Number** | **No Sleep Problems group** | **Sleep Problems group** | **noSP – SP** |
| --- | --- | --- | --- |
| Baseline | 28.8 (0.342; 28.1 - 29.4) | 30.5 (0.356; 29.8 - 31.2) | **-1.75 (0.45; 0.0001)** |
| Month 6 | 23.5 (0.415; 22.7 - 24.3) | 30.9 (0.429; 30.1 - 31.8) | **-7.4 (0.57; <0.0001)** |
| Month 9 | 23.4 (0.416; 22.6 - 24.2) | 30.4 (0.44; 29.5 - 31.2) | **-6.97 (0.57; <0.0001)** |
| Month 12 | 23.3 (0.483; 22.3 - 24.2) | 30.2 (0.509; 29.2 - 31.2) | **-6.94 (0.67; <0.0001)** |
| Month 15 | 22.7 (0.573; 21.6 - 23.9) | 30.6 (0.596; 29.4 - 31.8) | **-7.86 (0.8; <0.0001)** |
| Month 18 | 22.3 (0.669; 21 - 23.6) | 30.2 (0.713; 28.8 - 31.6) | **-7.85 (0.95; <0.0001)** |
| Month 21 | 20.9 (0.738; 19.5 - 22.4) | 31.2 (0.806; 29.6 - 32.8) | **-10.27 (1.07; <0.0001)** |
| Month 24 | 21 (0.813; 19.4 - 22.6) | 31.5 (0.962; 29.6 - 33.4) | **-10.52 (1.24; <0.0001)** |
| Month 27 | 20.3 (0.967; 18.4 - 22.2) | 29.8 (1.01; 27.9 - 31.8) | **-9.55 (1.38; <0.0001)** |
| Month 30 | 19.6 (1.138; 17.4 - 21.8) | 29.7 (1.113; 27.5 - 31.9) | **-10.12 (1.57; <0.0001)** |
| Month 33 | 20 (1.248; 17.5 - 22.4) | 32.1 (1.273; 29.6 - 34.5) | **-12.09 (1.77; <0.0001)** |
| Month 36 | 21.5 (1.199; 19.1 - 23.8) | 35.6 (1.535; 32.6 - 38.6) | **-14.14 (1.93; <0.0001)** |
| Month 36 - Baseline | -7.32 (1.21; <0.0001) | 5.07 (1.54; 0.001) | na |

**Table S6: LS Means (SE; 95% CI) for Combinatorial Receptive Language measured by the MSEC subscale when controlling for the health score. The difference between no Sleep Problems and Sleep Problems (noSP – SP) is presented as: LS Mean (SE; P-value). The negative noSP – SP difference indicates that the SP group had higher score and therefore more severe symptoms.**

| **Visit Number** | **No Sleep Problems group** | **Sleep Problems group** | **noSP – SP** |
| --- | --- | --- | --- |
| Baseline | 31.3 (0.319; 30.6 - 31.9) | 31 (0.318; 30.4 - 31.6) | 0.24 (0.31; 0.4244) |
| Month 6 | 29.2 (0.356; 28.5 - 29.9) | 29.5 (0.347; 28.8 - 30.1) | -0.24 (0.38; 0.5256) |
| Month 9 | 28.8 (0.357; 28.1 - 29.5) | 29.3 (0.353; 28.6 - 29.9) | -0.5 (0.39; 0.192) |
| Month 12 | 28.1 (0.39; 27.4 - 28.9) | 28.6 (0.393; 27.8 - 29.4) | -0.44 (0.45; 0.32) |
| Month 15 | 27.3 (0.439; 26.4 - 28.1) | 28.8 (0.436; 27.9 - 29.6) | **-1.51 (0.53; 0.0046)** |
| Month 18 | 26.7 (0.493; 25.7 - 27.7) | 28.3 (0.51; 27.3 - 29.3) | **-1.57 (0.63; 0.0123)** |
| Month 21 | 25.7 (0.536; 24.7 - 26.8) | 27.8 (0.567; 26.6 - 28.9) | **-2.01 (0.71; 0.0044)** |
| Month 24 | 25.2 (0.576; 24.1 - 26.3) | 26.6 (0.659; 25.3 - 27.9) | -1.36 (0.81; 0.0936) |
| Month 27 | 24 (0.67; 22.7 - 25.3) | 25.7 (0.692; 24.4 - 27.1) | -1.73 (0.9; 0.0562) |
| Month 30 | 24.3 (0.776; 22.7 - 25.8) | 25.5 (0.753; 24.1 - 27) | -1.28 (1.03; 0.2131) |
| Month 33 | 24.4 (0.842; 22.7 - 26) | 24.6 (0.851; 23 - 26.3) | -0.26 (1.15; 0.8207) |
| Month 36 | 22.6 (0.811; 21 - 24.1) | 25.1 (1.013; 23.1 - 27.1) | **-2.54 (1.26; 0.0437)** |
| Month 36 - Baseline | -8.71 (0.79; <0.0001) | -5.92 (0.99; <0.0001) | na |

**Table S7: LS Means (SE; 95% CI) for Expressive Language measured by the Subscale 1 of ATEC when controlling for the health score. The difference between no Sleep Problems and Sleep Problems (noSP – SP) is presented as: LS Mean (SE; P-value). The negative noSP – SP difference indicates that the SP group had higher score and therefore more severe symptoms.**

| **Visit Number** | **No Sleep Problems group** | **Sleep Problems group** | **noSP – SP** |
| --- | --- | --- | --- |
| Baseline | 18.2 (0.245; 17.73 - 18.7) | 17.8 (0.244; 17.37 - 18.3) | 0.36 (0.24; 0.1285) |
| Month 6 | 16 (0.272; 15.45 - 16.5) | 16.1 (0.266; 15.6 - 16.6) | -0.13 (0.29; 0.6499) |
| Month 9 | 15.1 (0.273; 14.58 - 15.7) | 15.6 (0.27; 15.02 - 16.1) | -0.43 (0.3; 0.143) |
| Month 12 | 14.7 (0.297; 14.15 - 15.3) | 15.2 (0.299; 14.6 - 15.8) | -0.45 (0.34; 0.1891) |
| Month 15 | 14 (0.334; 13.34 - 14.6) | 14.2 (0.331; 13.58 - 14.9) | -0.24 (0.4; 0.5579) |
| Month 18 | 12.8 (0.374; 12.1 - 13.6) | 13.9 (0.386; 13.15 - 14.7) | **-1.07 (0.47; 0.0241)** |
| Month 21 | 12.5 (0.406; 11.69 - 13.3) | 13.5 (0.428; 12.62 - 14.3) | -0.97 (0.53; 0.0677) |
| Month 24 | 12.3 (0.435; 11.46 - 13.2) | 13 (0.497; 12.05 - 14) | -0.71 (0.61; 0.2453) |
| Month 27 | 12.1 (0.504; 11.09 - 13.1) | 12.7 (0.522; 11.68 - 13.7) | -0.63 (0.68; 0.3553) |
| Month 30 | 10.4 (0.583; 9.28 - 11.6) | 12.6 (0.567; 11.45 - 13.7) | **-2.13 (0.77; 0.0057)** |
| Month 33 | 11.1 (0.633; 9.91 - 12.4) | 11.1 (0.64; 9.83 - 12.3) | 0.06 (0.87; 0.9414) |
| Month 36 | 10.6 (0.61; 9.39 - 11.8) | 11.8 (0.761; 10.32 - 13.3) | -1.23 (0.94; 0.1935) |
| Month 36 - Baseline | -7.62 (0.59; <0.0001) | -6.03 (0.74; <0.0001) | na |

**Table S8: LS Means (SE; 95% CI) for Sociability measured by the Subscale 2 of ATEC when controlling for the health score. The difference between no Sleep Problems and Sleep Problems (noSP – SP) is presented as: LS Mean (SE; P-value). The negative noSP – SP difference indicates that the SP group had higher score and therefore more severe symptoms.**

| **Visit Number** | **No Sleep Problems group** | **Sleep Problems group** | **noSP – SP** |
| --- | --- | --- | --- |
| Baseline | 18.3 (0.315; 17.7 - 18.9) | 17.6 (0.313; 17 - 18.2) | 0.69 (0.3; 0.0221) |
| Month 6 | 17.1 (0.353; 16.4 - 17.8) | 17 (0.343; 16.3 - 17.6) | 0.16 (0.38; 0.668) |
| Month 9 | 16.4 (0.353; 15.7 - 17.1) | 16.4 (0.349; 15.7 - 17.1) | 0 (0.38; 0.9929) |
| Month 12 | 16.1 (0.387; 15.3 - 16.9) | 16.6 (0.39; 15.8 - 17.3) | -0.47 (0.45; 0.2891) |
| Month 15 | 16.4 (0.438; 15.5 - 17.2) | 15.8 (0.435; 14.9 - 16.6) | 0.58 (0.53; 0.2793) |
| Month 18 | 15.8 (0.493; 14.8 - 16.7) | 16.4 (0.51; 15.4 - 17.5) | -0.68 (0.63; 0.2839) |
| Month 21 | 16.1 (0.537; 15.1 - 17.2) | 16.5 (0.568; 15.4 - 17.6) | -0.38 (0.71; 0.5884) |
| Month 24 | 15.9 (0.578; 14.7 - 17) | 16.1 (0.662; 14.8 - 17.4) | -0.2 (0.82; 0.8031) |
| Month 27 | 15.6 (0.673; 14.3 - 16.9) | 15.2 (0.696; 13.9 - 16.6) | 0.38 (0.91; 0.6791) |
| Month 30 | 16.8 (0.782; 15.2 - 18.3) | 15.2 (0.758; 13.7 - 16.6) | 1.62 (1.03; 0.1188) |
| Month 33 | 16.3 (0.849; 14.6 - 18) | 16.2 (0.858; 14.5 - 17.9) | 0.1 (1.16; 0.9324) |
| Month 36 | 15.1 (0.818; 13.5 - 16.7) | 15.4 (1.023; 13.4 - 17.4) | -0.3 (1.27; 0.812) |
| Month 36 - Baseline | -3.19 (0.79; <0.0001) | -2.19 (1.01; 0.0292) | na |

**Table S9: LS Means (SE; 95% CI) for Sensory Awareness measured by the Subscale 3 of ATEC when controlling for the health score. The difference between no Sleep Problems and Sleep Problems (noSP – SP) is presented as: LS Mean (SE; P-value). The negative noSP – SP difference indicates that the SP group had higher score and therefore more severe symptoms.**

| **Visit Number** | **No Sleep Problems group** | **Sleep Problems group** | **noSP – SP** |
| --- | --- | --- | --- |
| Baseline | 17.9 (0.274; 17.3 - 18.4) | 17.5 (0.273; 17 - 18.1) | 0.32 (0.26; 0.2256) |
| Month 6 | 17 (0.306; 16.4 - 17.6) | 16.9 (0.298; 16.3 - 17.5) | 0.06 (0.33; 0.8587) |
| Month 9 | 16.4 (0.307; 15.8 - 17) | 16.6 (0.304; 16 - 17.2) | -0.23 (0.33; 0.4837) |
| Month 12 | 16.1 (0.336; 15.5 - 16.8) | 16.2 (0.339; 15.5 - 16.9) | -0.08 (0.39; 0.8332) |
| Month 15 | 15.8 (0.379; 15 - 16.5) | 15.9 (0.377; 15.1 - 16.6) | -0.11 (0.46; 0.8078) |
| Month 18 | 15.4 (0.427; 14.6 - 16.2) | 16.2 (0.442; 15.4 - 17.1) | -0.85 (0.55; 0.1178) |
| Month 21 | 14.7 (0.465; 13.8 - 15.6) | 15.7 (0.492; 14.7 - 16.7) | -0.99 (0.61; 0.108) |
| Month 24 | 15.2 (0.5; 14.2 - 16.2) | 16.2 (0.573; 15.1 - 17.3) | -1.03 (0.71; 0.1459) |
| Month 27 | 15 (0.583; 13.9 - 16.2) | 14.6 (0.603; 13.4 - 15.8) | 0.47 (0.79; 0.5539) |
| Month 30 | 15 (0.676; 13.7 - 16.3) | 15.2 (0.656; 13.9 - 16.4) | -0.16 (0.89; 0.856) |
| Month 33 | 14.9 (0.734; 13.5 - 16.4) | 15.9 (0.743; 14.4 - 17.4) | -0.96 (1.01; 0.3423) |
| Month 36 | 13.7 (0.707; 12.3 - 15.1) | 14.3 (0.885; 12.6 - 16) | -0.61 (1.1; 0.5765) |
| Month 36 - Baseline | -4.17 (0.69; <0.0001) | -3.24 (0.87; 0.0002) | na |
